# The “Diabetes in Primary Care – Improving Classification” (DePICtion) study: a feasibility study to determine the utility and acceptability of implementing diabetes classification tools in primary care

**DOI:** 10.64898/2026.10.20.26366000

**Authors:** Beverley M Shields, Katherine G Young, Maddy Geen, Anita Hill, Charlotte Lee, Robin Humphreys, Clare Timblin, Kevin Colclough, John Campbell, Willie Hamilton, Philip Evans, Angus Jones, Andrew Hattersley, Katrina Turner, Nadeem Qureshi

## Abstract

**Introduction:** There are several subtypes of diabetes, including type1, type 2, and a genetic form called MODY. Classification can be challenging, particularly in young people, with up to 15% of people misclassified with the wrong subtype, resulting in inappropriate treatment. We assessed the feasibility of embedding a decision support tool (the DePICtion tool) in GP records to flag patients at risk of misclassification for further investigation and biomarker testing.

**Research Design & Methods:** The DePICtion tool was run in 11 UK GP practices. The tool generated a list of patients at high probability of being misclassified for review by practice staff. After review, practices returned a follow-up data form, summarising the actions taken for these patients.

**Results:** Overall, 7% (237/3501) of patients were flagged as potentially misclassified. Of 203 with follow-up data, 28 (14%) records had coding errors and 2 patients had died, leaving 173 (85%) suitable for biomarker testing to confirm the diagnosis. Around half (88/173) of these patients received new biomarker testing or had previous testing on the system (73 complete, 15 partial or unknown results). 17% (30/173) of patients declined/did not respond. For the remaining 55, the practice reason for not testing did not rule out an alternative diagnosis in the majority of cases (n=44). Among the 73 patients with definitive results, nearly 1 in 5 (18%; 13) changed diagnosis (2 diagnosed with MODY, 6 from type 1 to type 2, 5 from type 2 to type 1).

**Conclusions:** The DePICtion tool identified people with diabetes that had been misclassified. However, many people flagged as potentially misclassified did not have classification biomarker testing. Better data recording and greater engagement with secondary care could improve uptake of testing to identify more misclassified patients and optimise their diabetes care.

**Key messages:** *What is already known on this topic:* Classification of diabetes subtypes can be challenging, and misclassification is common in clinical practice. Classification algorithms have been developed, but their utility in primary care is not known.

*What this study adds:* We carried out a feasibility study to determine the utility and acceptability of running the diabetes classification algorithms as automated decision support tools in 11 GP practices. We found that the algorithms did identify patients at high risk of misclassification, but that lack of specialist input and GP time were barriers to wider uptake.

*How this study might affect research, practice or policy:* In line with the NHS 10 year plan, there is a strong drive to move from analogue to digital, but implementation of statistical models into clinical practice needs robust studies to ensure their translation is useful in real-world care. This study provided key data to inform future steps towards large-scale roll out of this approach to ensure misclassified patients are appropriately identified and they can obtain the right treatment and care for their diabetes.

## Introduction

Diabetes is a heterogeneous disease classified into several different subtypes, which require distinct treatment and management. Type 1 diabetes is characterised by severe insulin deficiency requiring daily insulin injections, whereas in Type 2 diabetes, endogenous insulin secretion is maintained so these patients can largely be treated with behavioural change and tablets^1^. There are also rarer genetic subtypes, such as Maturity Onset Diabetes of the Young (MODY), where in most cases patients require either no pharmacological treatment or can achieve excellent glycaemic control with sulphonylurea tablets^2^.

Classification of diabetes is challenging, particularly in young adults, as clinical features often overlap. Consequently, up to 15% of adults treated with insulin for their diabetes^3–5^ and up to 80% of MODY cases^6^ are estimated to be misdiagnosed, resulting in many patients receiving inappropriate treatment and care. Biomarker testing can help determine the correct diagnosis, with C-peptide and/or islet autoantibodies aiding discrimination of Type 1 from Type 2 diabetes^7^ ^8^, and genetic testing can confirm MODY^2^. However, these tests are rarely performed in primary care, where most people with diabetes are managed.

Clinical decision support tools^9^, are being increasingly embedded in primary care electronic health records and could provide an automated approach to determine which patients with diabetes may be at risk of misclassification, prompting further investigation and biomarker testing to confirm their diagnosis. Two validated algorithms have been developed that are highly predictive for discriminating diabetes subtypes: a “Type 1/Type 2 calculator” for adults diagnosed aged 18-50^10^ ^11^, and a “MODY calculator”^12^ for people with diabetes diagnosed under age 35. These algorithms are based on routine clinical features, so could be run as automated searches on electronic healthcare records to flag patients at high probability of MODY or high risk of misclassification of Type 1 or Type 2.

We aimed to determine the feasibility of running these algorithms, as a clinical decision support tool (the DePICtion tool), in primary care electronic healthcare records to identify patients with diabetes who may have been misclassified.

## Methods

### Study population

We purposefully recruited GP practices from Plymouth and Greater Nottingham areas in the UK, serving populations with diverse levels of deprivation and proportions of ethnic minorities. Practice ethnicity breakdown and deprivation deciles were determined through the Department of Health and Social Care National General Practice Profiles (https://fingertips.phe.org.uk/profile/general-practice), with deprivation based on the Index of Multiple Deprivation 2025. Practices were approached through the South West Peninsula and East Midlands Local Clinical Research Networks and were eligible if they used the TPP (SystmOne) GP system and referred to Plymouth or Nottingham hospitals (where secondary care teams were aware of this study in case of increased referrals). We aimed to recruit sufficient practices to test the tool on ∼3000 eligible diabetes patients (see Supplementary Methods), so targeted practices with a list size of >10,000 patients, aiming for 10-12 practices in total.

### The DePICtion tool

The “**D**iab**e**tes in **P**rimary Care – **I**mproving **C**lassifica**tion**” (DePICtion) tool applied the algorithms (regression equations) behind the MODY^12^ and Type 1/Type 2^10^ ^11^ calculators on data automatically extracted from primary care records.

We developed a specification for extracting relevant data from the GP system, along with validation rules to remove erroneous values, based on previous work in UK primary care data (CPRD)^13^. This specification determined the database searches applied to the GP system to extract appropriate records for young adult patients diagnosed with diabetes (defined by a proxy of earliest diabetes code, excluding gestational diabetes) aged 50 years or under, in line with the original calculators.

Variables necessary for the Type 1/Type 2 and MODY calculator equations were extracted using clinical code lists and included sex, age, initial and current treatment (defined based on earliest and latest prescriptions for diabetes glucose-lowering medications), recorded family history of diabetes, BMI (latest available since diagnosis), HbA1c (latest available since diagnosis) and lipids (HDL, cholesterol, triglycerides; all latest available since diagnosis).

After applying the algorithms (model regression equations)^11^ ^12^ on the extracted data, the tool produced lists of patients at high probability of misclassification, based on pre-determined criteria (see below) for a nominated member of the general practice team to review. The search also identified patients where variables had not been coded (identified as “missing cases”).

### Flagging patients at high probability of MODY

The DePICtion tool calculated probability of MODY for all patients diagnosed with diabetes aged 1-35 (the valid age range for the calculator^12^). NHS Genomic Testing criteria for monogenic diabetes recommend genetic testing at >20% probability for patients not insulin treated, or >10% for patients treated with insulin^14^. As this was an initial feasibility study and the MODY calculator had not been assessed in primary care previously, to make this manageable at the practice level, we used a higher probability (>50%), estimated to lead to up to 3 patients for a practice of size 10,000 patients.

### Misclassification of more common forms of diabetes

The Type 1/Type 2 calculator equations^11^ were applied to all insulin treated patients diagnosed aged 18-50 to determine probability of type 1 diabetes (compared to type 2). Patients were flagged as potentially misclassified if:

1. Patients were coded on the GP system as type 1 diabetes (based on latest code), but had a type 1 diabetes probability <10% according to the tool (indicating possible type 2 diabetes).
2. Patients were coded on the GP system as type 2 diabetes (based on latest code), but had a type 1 diabetes probability >50% according to the tool (indicating possible type 1 diabetes).

The probability thresholds for indicating misclassification were established from previous work in UK Biobank assessing thresholds against genetically defined diabetes subtype^15^, and analysis in CPRD determining a manageable workload for practices^13^.

In addition, we also flagged patients coded as type 2 who required insulin treatment within 3 years from diagnosis, based on previous research demonstrating high rates of misclassification in this group^4^. Time to insulin was determined from the difference between the earliest diabetes code (proxy for age at diagnosis) and earliest insulin prescription (for insulin start date).

### The DePICtion tool outputs

The tool’s main output was a colour-coded summary page (see Supplementary Figure 1 for an example) detailing the numbers of patients found at high probability of MODY or misclassification of type 1/type 2 (coloured red) and numbers with missing information or coding errors (coloured yellow or orange) (see Supplementary Table 1 for full details). Practice staff could click on each category to see lists of individual patients, and then select a particular patient to review a summary of their GP record.

### Implementation of the tool

Each recruited practice had a site initiation session, comprising education from members of the study team (including a clinician) on diabetes misclassification and training on how to run the DePICtion tool and review the output. The session also provided an overview of the best practice guidelines on biomarker testing for classification of diabetes subtypes, including details of NHS genomic testing guidelines for MODY (monogenic diabetes)^14^, NICE guidelines for diabetes classification in adults^8^, and guidance on the use of C-peptide and islet autoantibody testing to confirm type 1 or type 2 diabetes^7^ ^16^.

Practices downloaded and ran the tool according to written instructions, with remote support from the research team and the tool developer (PRIMIS) if needed. After running the tool, practices returned a copy of the colour-coded summary page to the central research team so they had a record of the numbers flagged and to check that this was in line with what was expected. As this was a pragmatic study, general practices were advised to use their own discretion in terms of what further investigations to perform following running of the tool, based on current guidelines.

### Analysis

#### Collation and analysis of data collection sheets

By return of email, practices confirmed the dates they ran the tool and reviewed the records. At the end of implementation, practices returned a study follow-up data form to the research team providing anonymised details of the actions taken for each patient flagged by the tool, including coding errors found, contact with the patient, any biomarker testing carried out, referral to secondary care, and change in diagnosis and/or treatment, if applicable. A free text comments box was provided for any further detail. Given challenges in workload, they were asked to prioritise providing details for those in the red categories.

The data collection forms were compared to the initial summary sheets to check the numbers matched up. In addition, we asked the Exeter Genomics Laboratory (national diagnostic centre for MODY testing) to confirm the number of referrals from participating practices to cross-check and provide supplementary data in case practices did not provide follow-up information. Where there were discrepancies, practices were emailed to resolve issues relating to missing or additional data.

We coded each patient on the follow-up data form into a broad category for analysis, relating to whether they received biomarker testing or not, with further subcategories either based on the result of testing, or the reason why testing was not carried out (See Supplementary table 2 for the full list of categories). There was wide variability in how practice staff completed the form, with some filling in all boxes, but most only providing free text comments. Therefore, as the majority of coding was subjective based on free text comments, each case was classified by two authors (BS and AH), their classifications were compared, and discrepancies resolved through discussion.

As this was a feasibility study, analysis was largely descriptive, summarising the actions taken following running the tool.

## Results

103 Practices were approached via the NIHR regional delivery teams in South West Peninsula (13) & East Midlands (90). Disparity in numbers was due to a more general approach by East Midlands where invitations were not targeted at required list size or local secondary hospital. Two sites declined the expression of interest invitation, whilst 22 sites returned an expression of interest, with a total of 12 sites recruited to the study (6 South West, 6 East Midlands). Of the 10 not recruited, two had referral pathways to hospitals other than Plymouth or Nottingham, three had small list sizes (<10,000 patients), one did not respond to further contact, one had a site initiation visit but did not complete the study set up, and the remaining three were kept on a reserve list, but not recruited as we had reached capacity.

Demographics of the recruited practices are shown in Table 1 (one site represented three practices as had an over-arching database). The mean practice population size was 15,282. Practices included patients from diverse socioeconomic backgrounds and 2-37% of the patient population from ethnic minorities.

**Table 1.** Demographics and summary data for recruited practices in the DePICtion study.

| GP Practice Site | Population Size | Practice deprivation Score decile<sup>†</sup> | Practice Ethnicity Mix |  |  |  |  | Practice % Male |
| --- | --- | --- | --- | --- | --- | --- | --- | --- |
|  |  |  | White | Asian | Black | Mixed | Other |  |
| 1 | 16945 | 5 | 98% | 2% Non-white** |  |  |  | 49.5% |
| 2 | 15855 | 10 | 89.1% | 5.4% | 1.6% | 2.8% | 1.1% | 48.5% |
| 3 | 13376 | 3 | 70.2% | 9.5% | 11.4% | 6.9% | 2% | 48.6% |
| 4* | 43350 | 9 | 97.9% | 2.1% Non-white** |  |  |  | 48.3% |
| 5 | 10143 | 1 | 77.6% | 5.4% | 9.5% | 6.1% | 1.4% | 48% |
| 6 | 19400 | 4 | 92.8% | 2.8% | 1.3% | 2% | 1.1% | 51.3% |
| 7 | 26607 | 10 | 96.3% | 1.7% | - | 2% | - | 48.8% |
| 8 | 13572 | 9 | 85.4% | 7.1% | 2.2% | 3.3% | 2% | 50.5% |
| 9 | 12610 | 6 | 62.8% | 18.6% | 8.9% | 5.6% | 4.1% | 53.3% |
| 10<sup>‡</sup> | 11500 | 8 | 95.5% | 1.8% | - | 1.2% | 1.5% | 47.2% |

\*Site 4 was one over-arching database comprising 3 practices under one centre. † Index of Multiple Deprivation (IMD) 2025 decile derived from https://fingertips.phe.org.uk/profile/general-practice (1 = most deprived and 10 = least deprived). ‡ Practice recruited but did not run the tool. \*\*More detailed breakdown by ethnicity not available.

### Timeline for running decision support tool searches and actions completed

11 of the 12 practices ran the tool. There was considerable variability in how long it took them to run the tool, review the lists, and contact patients (total time ranging from 51-238 days; Supplementary Figure 2). In most cases, the tools were run by the practice managers, whilst patient records were mostly reviewed by GPs (7 practices) and/or nurses (5 practices) (Supplementary Table 3).

10 out of the 11 practices that ran the tool returned their follow-up data form. For the practice that did not return their form, we confirmed the numbers they referred for MODY testing with the national genetics laboratory, but were unable to determine any further details on the actions they took. Only two practices provided (limited) details on the yellow or orange categories, therefore, we only present summary results for the red (high priority) categories.

### Overall extent of misclassification identified by the DEPICTION tool (Figure 1)

Figure 1 shows follow-up of the 237/3501(7%) eligible patients flagged by the tool as potentially misclassified. Of 203 with follow up data available, 28 were found to be coding errors in the primary care electronic health records (9 wrong ages at diagnosis, 19 wrong diabetes type/other known cause), and 2 patients had died, leaving 173 suitable for further biomarker testing. Around half of these patients (88/173 (51%)) received biomarker testing (33 had previous results, 55 new testing). For the remaining 55 patients, in the majority of cases (n=44), the practice reason for not testing did not rule out an alternative diagnosis.

**Figure 1.**
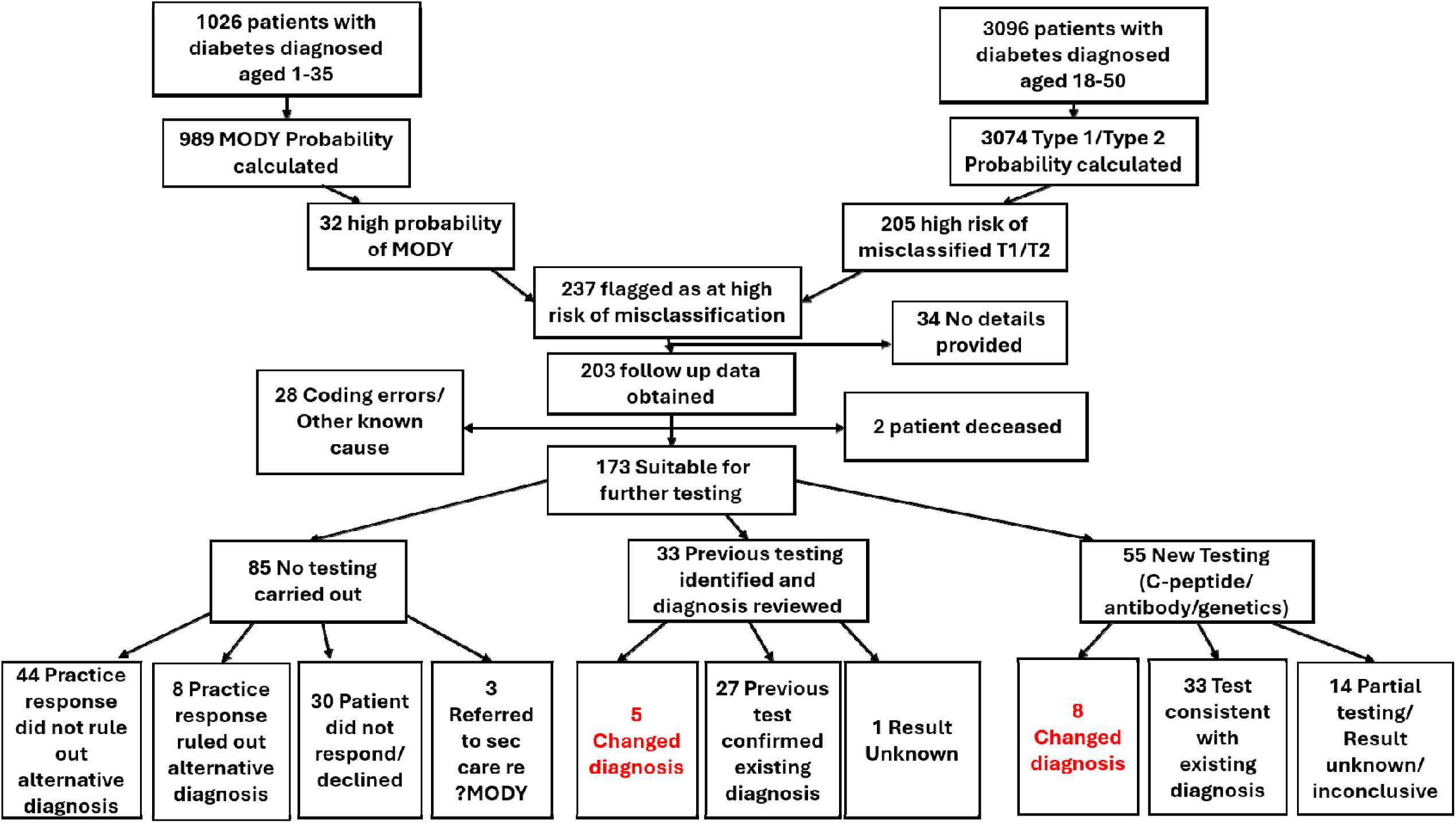
Flow diagram providing an overall summary of the patients flagged and actions taken across the 11 practices who ran the DePICtion tool.

Of the 73 patients with complete classification biomarker testing (excluding 15 with partial or unknown/inconclusive results), 13 patients (18%) changed diagnosis as a result of further investigation (Figure 1): 2 diagnosed with MODY, 6 changed from type 1 to type 2, and 5 from type 2 to type 1

Summary data by practice is presented in Supplementary Table 3.

### Identifying Patients for MODY genetic testing (Figure 2; Supplementary Table 4)

Across the 11 practices, 1026 patients were diagnosed with diabetes between the ages of 1 and 35 and hence were eligible for the MODY calculator. Of the 1026 eligible patients, 3 (0.3%) were already diagnosed with MODY and 989 had complete data (Figure 2; Supplementary Table 4).

**Figure 2.**
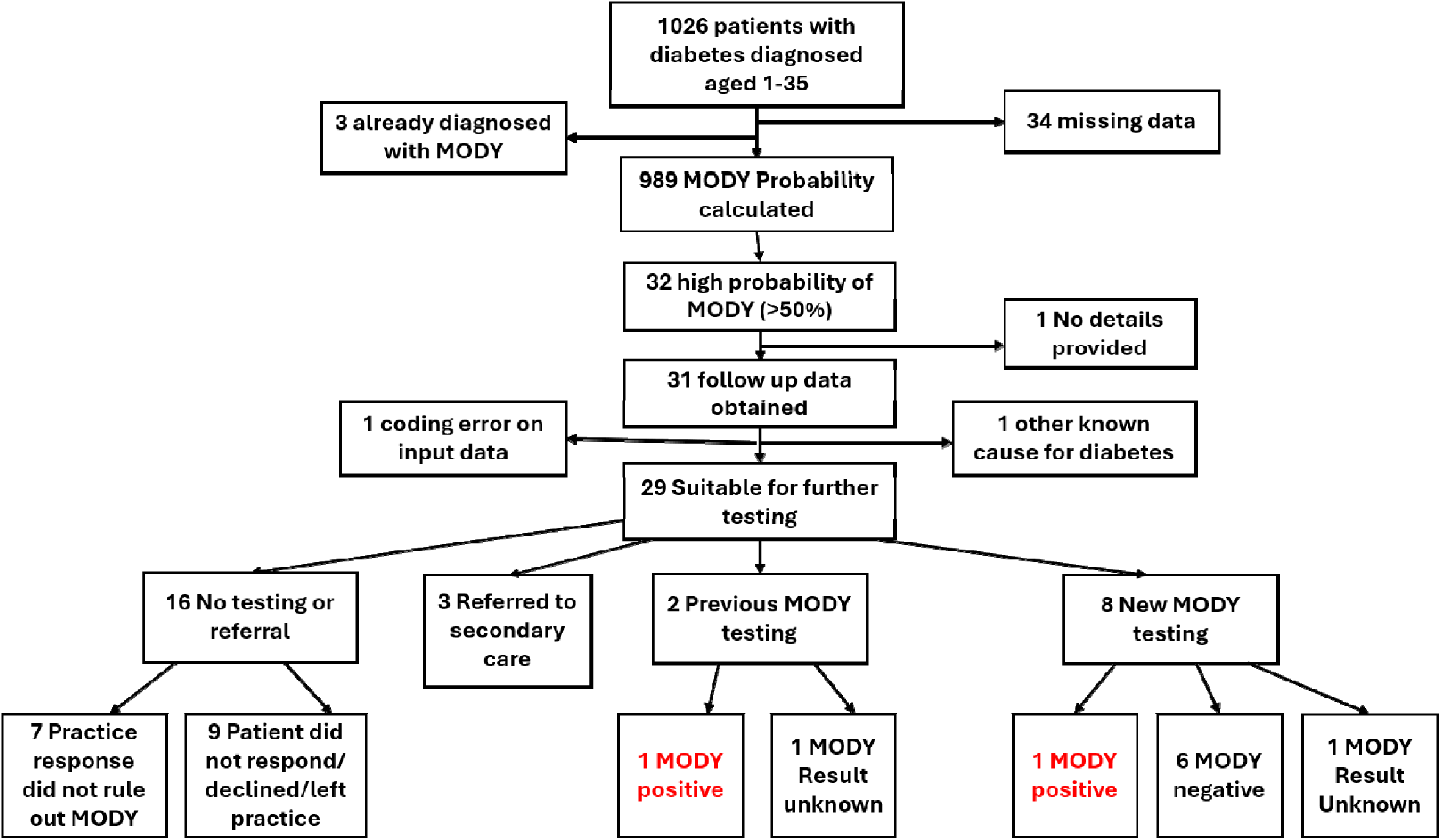
Flow diagram showing the follow up of patients flagged at high probability of MODY by the DePICtion tool.

32 patients were identified at high probability of MODY, and 29 had follow-up and were suitable for further testing, 2/29 had previous testing for MODY (one positive and recoded on the GP computer system, the other unclear (free text: *“Notes reviewed. Previous Cpep, GAD and MODY results. Already under secondary care”*)). Eight received new genetic testing for MODY, and three further patients were referred to secondary care for further review. Of 7 patients where the outcome of genetic testing was available 1 had tested positive.

In 16/29 (55%) cases suitable for testing, no action was taken. In 7 of these cases, the practice staff indicated they had not carried out testing, but their free text reasons did not rule out MODY (e.g. *“GAD and ZNTR8 negatikve[sic], insulin used for rescue therapy as symptomatic at time of diagnosis”; “Gestational diabetes”).* In the other 9 cases, the reason for not testing was due to patient factors (6 did not respond, 2 declined, 1 left the practice).

### Identifying Patients for Type 1/Type 2 biomarker testing

3096 patients were diagnosed with diabetes between the ages of 18 and 50, inclusive, and so met criteria for assessment against the Type 1/Type 2 diabetes classification algorithm (a mean of 281 per practice). Of 3096 eligible patients, 22(0.7%) had missing data and so their probability could not be calculated (Figure 3; Supplementary table 5).

**Figure 3.**
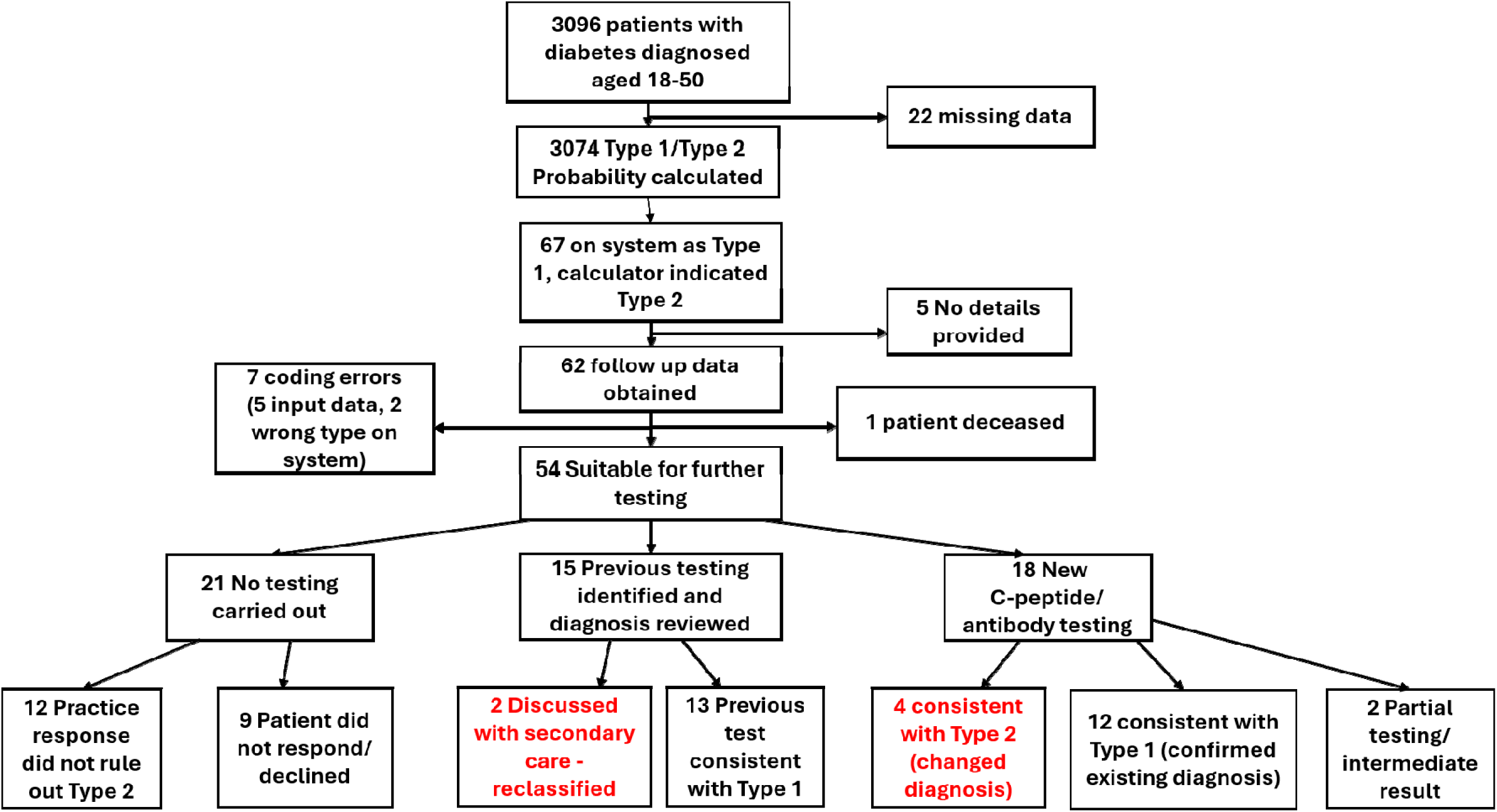
Flow diagram showing the follow up of patients coded as Type 1 on the GP system but the DePICtion tool flagged at low probability of Type 1 (indicating Type 2).

#### a) Patients coded as Type 1 but flagged as possible Type 2 (*Figure 3*)

A total of 67 patients were coded as type 1 on the GP system, but the DePICtion tool indicated they may have type 2, and 54 of these had follow-up data and were suitable for further testing. Of these, 33/54 (61%) had new or previous biomarker testing, resulting in 6/33 of those tested (18%) changing diagnosis: 4 based on new C-peptide or antibody testing and 2 after review of previous test results and discussion with secondary care. Anecdotally, many of the previous biomarker test results were reported to be located in free text in the scanned hospital letters rather than coded in the patient record. 21 patients had no testing (12 because this was not instigated by the practice, and 9 due to patients declining/not responding). In 9/12 cases where the practice decided not to test, the reason given was that the patient was “*already under secondary care*”. Other responses justified decisions by stating clinical symptoms indicated a type 1 diabetes diagnosis, but the symptoms reported were not sufficient alone to rule out type 2 (e.g. “*Type 1, slow onset*”, and “*10 day history of sx, ketones in urine, started on insulin immediately*”[sic]).

#### b) Patients coded as Type 2 but flagged as possible Type 1 (*Figure 4*)

Fewer type 2 patients were potentially misclassified, with 18 patients coded as type 2 on the GP records but flagged at high probability of type 1 (Figure 4). Of the 18 patients, 10 had follow-up data and were suitable for further testing. Two patients had previous high C-peptide results confirming their current diagnosis. New classification biomarker testing was only undertaken in one individual, where high C-peptide confirmed their existing diagnosis (type 2). In 4 cases, the practice staff did not instigate testing after reviewing the notes, with free text comments providing justification for excluding Type 1 diabetes in one case *(“5 years oral treatment prior to insulin”*), but not for the other 3. Three patients did not respond to contact from the practice.

**Figure 4.**
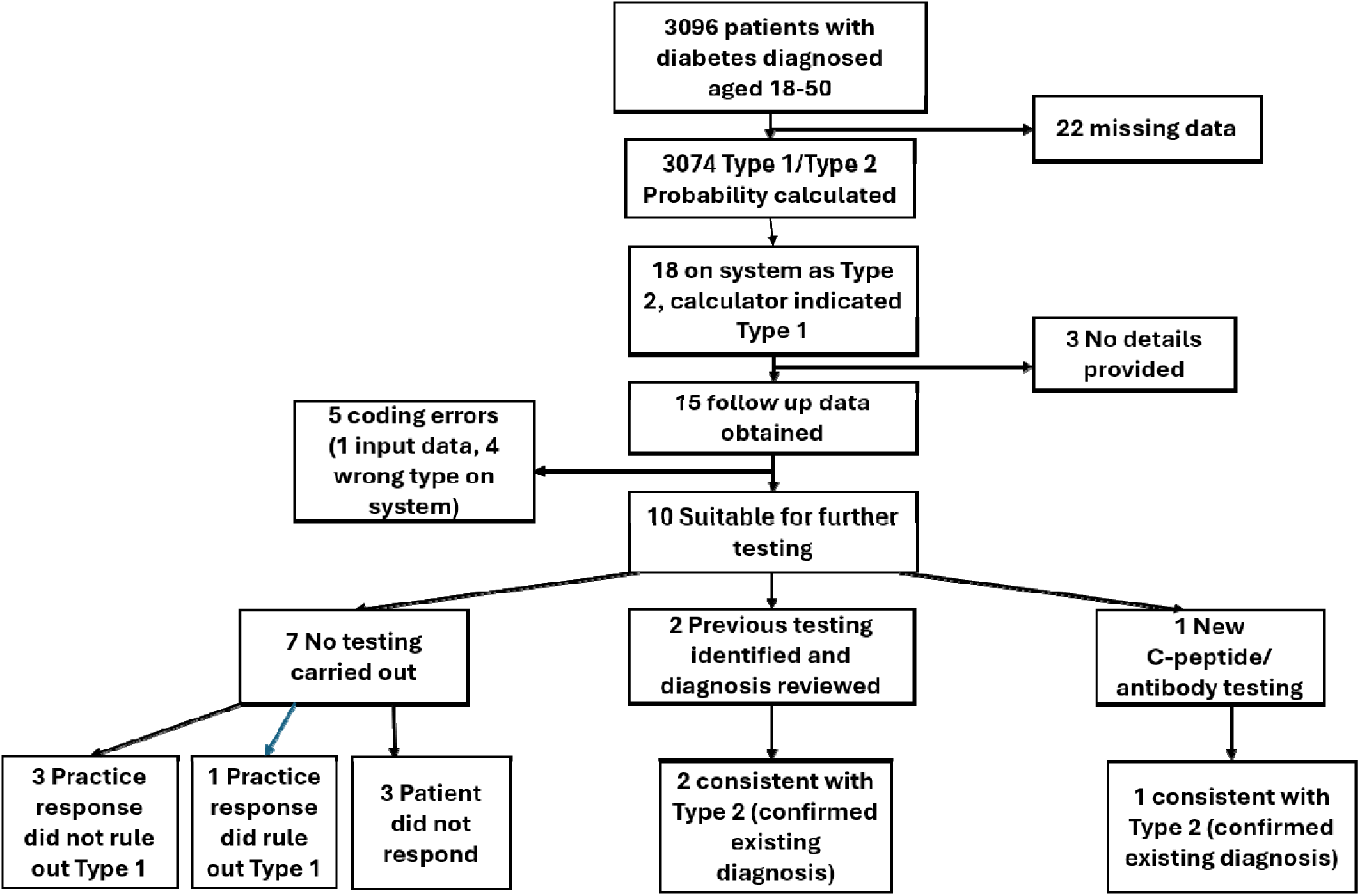
Flow diagram showing the follow up of patients coded as Type 2 on the GP system but the DePICtion tool flagged at high probability of Type 1.

#### c) Patients coded as Type 2 but rapidly progressed to insulin within 3 years (*Figure 5*)

A total of 120 patients were coded on the GP system as Type 2 but rapidly progressed to insulin within 3 years, indicative of Type 1 (Figure 5). Of these 120 patients, 80 had follow-up data and were suitable for further testing. Of these, 42/80 (53%) had either had previous testing or received new testing after using the tool. In 11 of these 42 cases (26%) testing was inconclusive (4 intermediate C-peptide results (200-600pmol/L) and 7 antibody negative but no C-peptide to rule out antibody negative Type 1). In those with conclusive test results (n=31), 5 (16%) were reclassified as type 1. In 9/80 cases (11%) the patient did not respond or declined further testing. In 29/80 cases (36%), the practice staff did not contact patients for further testing. For 7/29, responses from the practices highlighted clinical signs clearly ruling out Type 1 (e.g. patient stopped insulin), whilst in the other 22 cases reasons provided were insufficient to rule out Type 1 (e.g. *“HbA1c improved since starting GLP1”)*.

**Figure 5.**
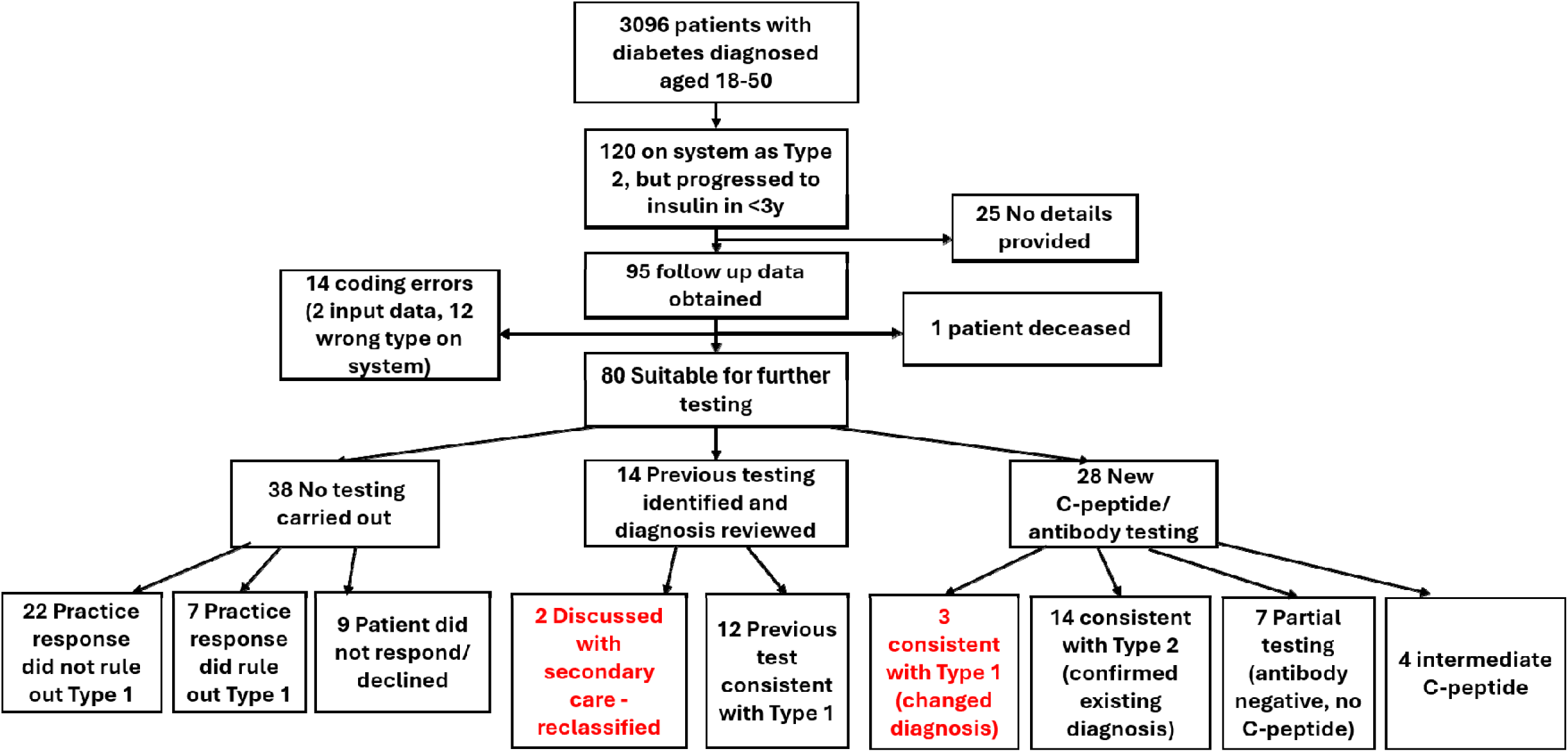
Flow diagram showing the follow up of patients coded as Type 2 on the GP system but progressing to insulin w thin 3 years indicative of Type 1 diabetes

## Discussion

### Summary

We showed that a diabetes classification decision support tool, that provides an automated search of GP records identified young-onset patients who had been given the incorrect diagnosis for their type of diabetes. By running the tool, 7% of people with diabetes diagnosed ≤50 years old (mean 22 per practice) were flagged for further assessment. Practice staff initiated or confirmed previous biomarker testing in 51% of eligible cases, and nearly 1 in 5 of those who completed testing had their diagnosis changed.

### Strengths and limitations

This is the first study to evaluate a decision support tool in primary care to identify patients with diabetes that may have been misclassified. The tool was easy to implement, and minimal missing data (∼3%) meant it could generate results for nearly all eligible patient records. The study was undertaken in practices with diverse deprivation scores and varying proportions of minority communities in the practice population.

To improve generalisability to routine care, no direct resource was provided for practices to implement findings from the tool (beyond modest reimbursement for research data collection). This may reflect realistic outcomes in routine care where additional resources may not be available, but meant with limited resources and other priorities, many patients identified as high risk by the tool were not followed up.

### Comparison with existing literature

Previous studies indicate approximately 7-15% of adults diagnosed with type 1 diabetes are misclassified^3^ ^4^ ^17^ ^18^, and around 5% of insulin-treated type 2^4^. The number that changed diagnosis in our study was much lower, but this is likely to represent a substantial underestimate of the true proportion of misclassified patients, as we used high probability thresholds for flagging patients to minimise GP workload and only half of those flagged received biomarker testing, with conclusive results available for only 42% (n=73). In addition, the proportion who had previous biomarker testing prior to running the tool (19%) was much higher than anticipated (<1% have C-peptide or islet autoantibody results in CPRD), meaning misclassified cases may have already been picked up prior to running the tool, and so the tool may be more beneficial in places where there has been lower uptake of testing previously. In previous research, largely from secondary care, around a third of those coded as type 2 but required insulin treatment within 3 years from diagnosis, were found to have type 1^4^, but in this study the proportion was much lower (12% of those tested), and may be due to a different mix of diabetes in secondary care.

Classification biomarker testing was not carried out in around half of cases flagged, due to a mix of GP and patient decisions. In the majority of cases, reasons provided did not rule out misclassification, often citing that the patient was “already in secondary care” (where misclassification is still common^3^) or by providing signs and symptoms that could still be consistent with the alternative diagnosis. In-depth interviews held with staff in the participating practices indicated GPs were often reluctant to question the diagnoses of patients under secondary care, and lacked confidence when making clinical decisions about misclassification^19^.

A significant proportion (17%) of patients declined testing or did not respond. This is consistent with other studies where GP record searches have identified individuals at increased risk of specific medical conditions but there was lack of patient engagement^20^ ^21^. As part of this feasibility study, interviews were held with patients that had been flagged by the tool^19^. These highlighted that people were most likely to want their diabetes consultant to be responsible for querying their diagnosis, and some perceived reclassification as a threat, particularly flagging the stigma associated with a diagnosis of Type 2 diabetes^19^.

The MODY calculator had not previously been tested in primary care, but it did identify patients that met NHS national criteria for genomic testing. Out of 7 cases that underwent new testing, only one case was positive, lower than we anticipated based on the >50% probability threshold, but numbers were small so it is hard to draw meaningful conclusions as wide uncertainty around estimates. We have recently developed an updated version of the model with improved recalibration^22^, specifically tested against UK primary healthcare records, so this newer improved model may yield better detection rates in future iterations.

### Implications for research and practice

The DePICtion tool did successfully determine patients suitable for classification biomarker testing and identified people who had been misdiagnosed or miscoded with the wrong type of diabetes, including patients meeting national NHS genomic testing criteria for MODY, rarely picked up in primary care routinely. In the current climate of managing the demand for genetic tests, further assessment of the testing criteria may be needed to identify the most cost-effective threshold to use in primary care.

Rollout across all UK general practices could lead to improvements in diabetes care. Previous research has shown that receiving the right diagnosis leads to both cost savings for the NHS, and major benefits for the patients, who receive more appropriate treatment and improved glucose control, with those who are able to stop insulin describing it as “life-changing”^3^ ^23–25^.

However, further work is needed to optimise the intervention before a pragmatic trial or service development project, as testing was only conducted in half of cases. This highlights the importance of conducting feasibility studies such as this, as the performance of statistical models developed in research settings may not translate directly into clinical care and it is important to identify real-world barriers that need to be addressed to enable their optimal use. As a result of this work, we have identified clear areas where we could look to improve how the tool is implemented for wider roll out to detect even more patients needing their diabetes subtype reviewed. In particular, enabling greater engagement with secondary care teams when reviewing patients may help improve uptake of testing, and supporting practices with appropriate education and resources would help boost primary care’s confidence to undertake further testing and amend the diabetes diagnosis when misclassified.

Key information was identified in free text in GP records and scanned hospital letters, not readily accessed by automated search/audit tools. Further administrative resources are needed to improve coding, which will be increasingly important as the NHS moves from analogue to digital in line with the NHS 10 year plan.

In conclusion, searching primary care records using the DePICtion tool identified people with diabetes who had been misclassified and/or miscoded. However, many people flagged as high risk of misclassification did not proceed to classification biomarker testing. Better data recording and greater engagement with secondary care, could improve uptake of testing to identify more misclassified patients and optimise their diabetes care.

### Funding

This study was funded by the National Institute for Health and Care Research (NIHR) School for Primary Care Research (SPCR) and supported by the NIHR Exeter Biomedical Research Centre. The views expressed are those of the author(s) and not necessarily those of the NIHR or the Department of Health and Social Care.

## Data Availability

All data produced in the present study are available upon reasonable request to the authors.

## Acknowledgements

We would like to thank the NIHR research delivery networks for supporting recruitment, and the primary care teams who participated in this project. Thanks to PRIMIS for providing the software for the decision support tool. We thank the Exeter PPIE group, patients and GPs in the Nottingham Deep end network, and the Indian Community Centre Association (ICCA) for providing their feedback on this project and its findings.

## Ethical approval

Ethical approval was granted by the NHS Health Research Authority (reference: 23/SC/0319).

