## Supplementary Material for "The “Diabetes in Primary Care – Improving Classification” (DePICtion) study: a feasibility study to determine the utility and acceptability of implementing diabetes classification tools in primary care"

Supplementary Methods

**Sample size calculation**

This study aims to provide estimates of prevalence of misclassification in primary care to inform the sample size for a full trial. We estimate for a practice of average list size, that there will be on average 12 misclassified patients (see below).

The original sample size was based on up to 20 general practices and an average list size of 7,900 based on a previous study6, covering an estimated population of 158,000 people. This would lead to an estimated 3318 patient records eligible for the Type 1/Type 2 calculator (7% of the population have diabetes1, and 30% of diabetes patients are diagnosed between 18 and 50 with a diagnosis of either Type 1 or Type 2 diabetes (from CPRD)), and 1549 for the MODY calculator (14% of people with diabetes are under the age of 35 (from CPRD)). Due to additional costs, we targeted fewer practices at higher populations to cover the same population.

This sample size would enable us to estimate the prevalence of MODY in general practice within a subgroup of young onset diabetes with tight confidence intervals. Based on previous prevalence estimates, we would estimate that 4% of the population in this age group will have MODY12. This would lead to an estimated 62 patients with MODY (with 95% confidence interval 48 to 79; an average of 3.2 patients with MODY per practice). We would anticipate that only 2 of these 62 will be already identified (only 0.18% of diabetes in this age range have a clinical code for MODY in an initial code search in CPRD data).

For the Type 1/Type 2 calculator, again this study should provide an adequate sample size to appropriately determine the rates of misclassification in primary care with tight confidence intervals. 14% of patients diagnosed with diabetes aged 18-50 have Type 1 diabetes24 (n=465) and based on cross-sectional data from running the algorithm in the Exeter-based DARE cohort (against gold-standard C-peptide defined classification outcomes) we estimate that 15% will be misclassified leading to a total of 70 patients across all practices (95% confidence interval 55 to 88; an average of 3.5 per practice). 86% of patients in this age range will have Type 2 (n=2853) and based on the same approach analysing the DARE cohort we estimate 4% will be misclassified, leading to a total of 114 patient across all practices (95% CI 94 to 136; an average of 5.7 per practice).

It is important to emphasise that these estimates are based on research studies with patients largely recruited from secondary care, and that a key aim of this work is to determine how appropriate they are in primary care.

Supplementary Table S1 Colour coding of categories of patients in the summary sheet of the DePICtion tool

| **Category** | **Criteria for patients flagged in this category** |
| --- | --- |
| Red (highest priority – high risk of misclassification) | High probability of MODY  High probability of Type 1 but coded as Type 2  Low probability of Type 1 (likely Type 2) but coded as Type 1  Coded as Type 2 but rapid progression to insulin within 3 years |
| Orange (medium priority – review records) | Missing coded family history data, but would be high probability if parent has diabetes  Coded as type 1 diabetes but on oral medication (except metformin) |
| Yellow (lower priority – likely coding errors) | Coded as Type 1 but no insulin prescriptions  Coded as Type 1 but no insulin within the first 5 years of diagnosis |
| White (optional audit) | Missing data |

Supplementary Table 2 – details of categorisation of the follow-up data form for GP responses to actions carried out for patients flagged at high risk of misclassification (red category) by the DePICtion tool.

**Categorisation of GP action**

No details

No test - Coding error (input data)

No test - Coding error (diabetes type)

No test - No response from patient

No test - Patient declined

No test - Patient left practice

No test - Referred to secondary care

No test – Patient deceased

No test - Other known cause

No test – GP response ruled out alternative diagnosis

No test – GP response did not rule out alternative diagnosis

Previous test – consistent with alternative diagnosis

Previous test – consistent with existing diagnosis

Previous test – result unknown/uncertain

New test – consistent with alternative diagnosis

New test – consistent with existing diagnosis

New test – result unknown/uncertain/intermediate/partial testing

Supplementary Table 3 - Summary data of reviewing and actions by practice after running the tool.

| **GP Practice** | **Non-White ethnicity (%)** | **Deprivation Decile (1=most deprived)** | **Staff who ran the tool** | **Staff who reviewed the records of patients flagged by the tool** | **Total eligible for the tool (n)** | **Flagged for testing (n (% of those eligible))** | **Total receiving testing – previous or new (n (% of those flagged)** | **No testing – patient declined/ did not respond (n (% of flagged)** | **No testing – GP did not initiate (n (% of those flagged)** |
| --- | --- | --- | --- | --- | --- | --- | --- | --- | --- |
| 1 | 1.90% | 5 | Practice Manager | 1 x Nurse | 326 | 24 (7.4%) | 12 (50%) | 5 (21%) | 5 (21%) |
| 2 | 10.80% | 10 | Practice Manager | 3 x GP | 316 | 22 (7.0%) | 10 (45%) | 1 (5%) | 5 (23%) |
| 3 | 28.50% | 3 | Practice Manager | 1 x GP | 378 | 24 (6.3%) | 16 (67%) | 5 (21%) | 2 (8%) |
| 4* | 2.10% | 9 | GP | 1 x GP, 3 x Nurse | 728 | 53 (7.3%) | 21 (40%) | 4 (8%) | 6 (11%) |
| 5 | 22.20% | 1 | ANP | 1 ANP, 1 Nurse | 290 | 25 (8.6%) | 3 (12%) | 3 (12%) | 11 (44%) |
| 6 | 5.10% | 4 | Practice Manager | 1 x GP, 1 x Nurse | 494 | 29 (5.9%) | 8 (28%) | 5 (17%) | 14 (48%) |
| 7 | 3.60% | 10 | Practice Manager | 1 x GP | 408 | 22 (5.4%) | 14 (64%) | 5 (23%) | 0 (0%) |
| 8 | 15.40% | 9 | Practice Manager | 1 x GP | 277 | 14 (5.1%) | 0 (0%) | 2 (14%) | 12 (86%) |
| 9† | 39.40% | 6 | Practice Manager | 1 x GP, 1 x Nurse | 284 | 24 (8.5%) | 4† | N/A† | N/A† |

†Practice 9 did not return their data collection form, but we know 4 received genetic testing from the numbers provided by the genomics laboratory; *Practice 4 comprised 3 practices but with one over-arching database. ANP=Advanced Nurse Practitioner

Supplementary Table 4. Summary data for recruited practices that ran the tool in the DePICtion study relating to potential MODY misclassification. * Practice 4 comprised 3 practices under one dataset (therefore means are for totals divided by 11 practices).

| **GP Practice** | **Population Size** | **N (%) with diabetes code** | **N (%) with diabetes eligible for MODY (age diag 1-35)** | **N (%) with missing data** | **>50% MODY Probability (n (%))**  (RED category) | **High MODY prob if FH positive (FH not known)**  (ORANGE category) | **Already diagnosed with MODY on system** |
| --- | --- | --- | --- | --- | --- | --- | --- |
| 1 | 16945 | 1204 (7.1%) | 85 (7%) | 6 | 2 | 0 | 0 |
| 2 | 15855 | 870 (5.5%) | 99 (11%) | 3 | 1 | 1 | 0 |
| 3 | 13376 | 877 (6.6%) | 89 (10%) | 2 | 5 | 1 | 0 |
| 4* | 43350 | 2487 (5.7%) | 240 (10%) | 6 | 8 | 5 | 0 |
| 5 | 10143 | 641 (6.3%) | 103 (16%) | 5 | 6 | 0 | 0 |
| 6 | 19400 | 1420 (7.3%) | 159 (11%) | 2 | 3 | 1 | 2 |
| 7 | 26607 | 1391 (5.2%) | 121 (9%) | 5 | 0 | 0 | 1 |
| 8 | 13572 | 824 (6.1%) | 75 (9%) | 2 | 2 | 0 | 0 |
| 9 | 12610 | 596 (4.7%) | 55 (9.2%) | 3 | 5 | 0 | 0 |
| **Mean** | **15623** | **937** | **93** | **3** | **3** | **1** | **0.4** |
| **Total** | **171858** | **10310** | **1026** | **34** | **32** | **8** | **3** |

Supplementary Table 5. Summary data for recruited practices that ran the tool in the DePICtion study relating to potential Type 1 and Type 2 misclassification. * Practice 4 comprised 3 practices under one dataset (therefore means are for totals divided by 11 practices).

| **GP Practice** | **Population Size** | **N (%) with diabetes code** | **N (%) with diabetes eligible for T1/T2 (age diag 18-50)** | **N (%) with missing data** | **GP code=T1, Model=T2**  (RED category) | **GP code=T2, Model=T1**  (RED category) | **GP code=T2, Insulin treated <3 years**  (RED category) |
| --- | --- | --- | --- | --- | --- | --- | --- |
| 1 | 16945 | 1204 (7.1%) | 287 (24%) | 2 | 8 | 2 | 12 |
| 2 | 15855 | 870 (5.5%) | 278 (32%) | 2 | 4 | 1 | 16 |
| 3 | 13376 | 877 (6.6%) | 347 (40%) | 2 | 6 | 1 | 12 |
| 4* | 43350 | 2487 (5.7%) | 622 (25%) | 7 | 20 | 4 | 21 |
| 5 | 10143 | 641 (6.3%) | 256 (40%) | 5 | 3 | 0 | 16 |
| 6 | 19400 | 1420 (7.3%) | 436 (31%) | 2 | 12 | 5 | 9 |
| 7 | 26607 | 1391 (5.2%) | 354 (25%) | 0 | 8 | 1 | 13 |
| 8 | 13572 | 824 (6.1%) | 251 (30%) | 1 | 2 | 1 | 9 |
| 9 | 12610 | 596 (4.7%) | 265 (45%) | 1 | 4 | 3 | 12 |
| **Mean** | **15623** | **937** | **281** | **2** | **6** | **2** | **11** |
| **Total** | **171858** | **10310** | **3096** | **22** | **67** | **18** | **120** |

Supplementary Figure 1 – Screenshot of the DePICtion tool summary page, with numbers provided showing the estimated average number of patients flagged based on a practice population of 10,000.

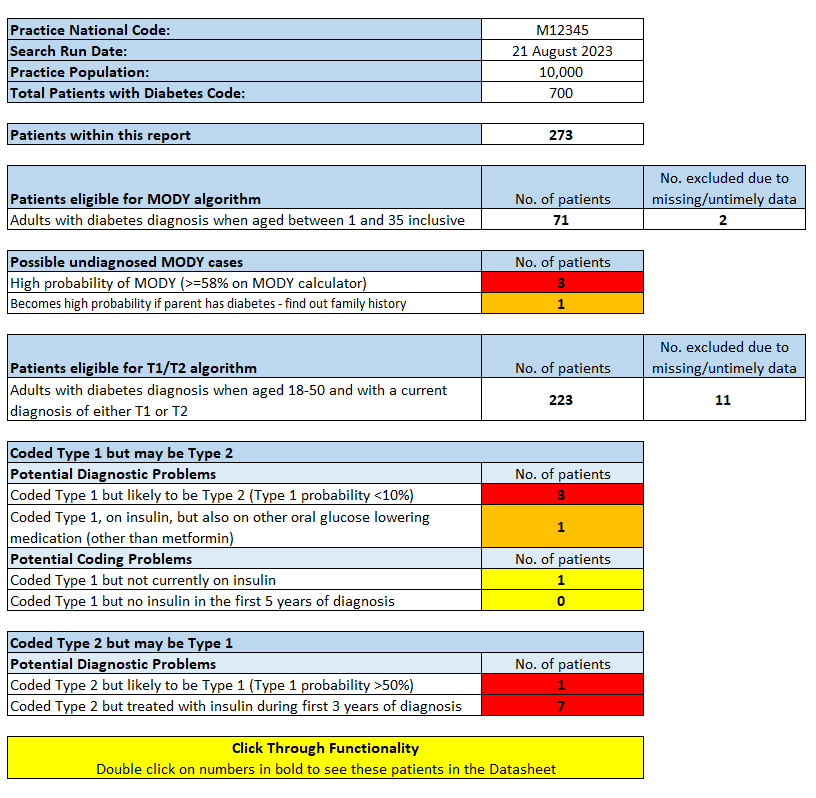

Supplementary Figure 2 – Timeline for each practice of time from site initiation visit to running the tool and reviewing patients. Practices 2, 3, 4, and 7 reviewed the list and contacted patients on the same day, so time from clinician review to patient contact is 0. *Practice 4 represents 3 practices in one.

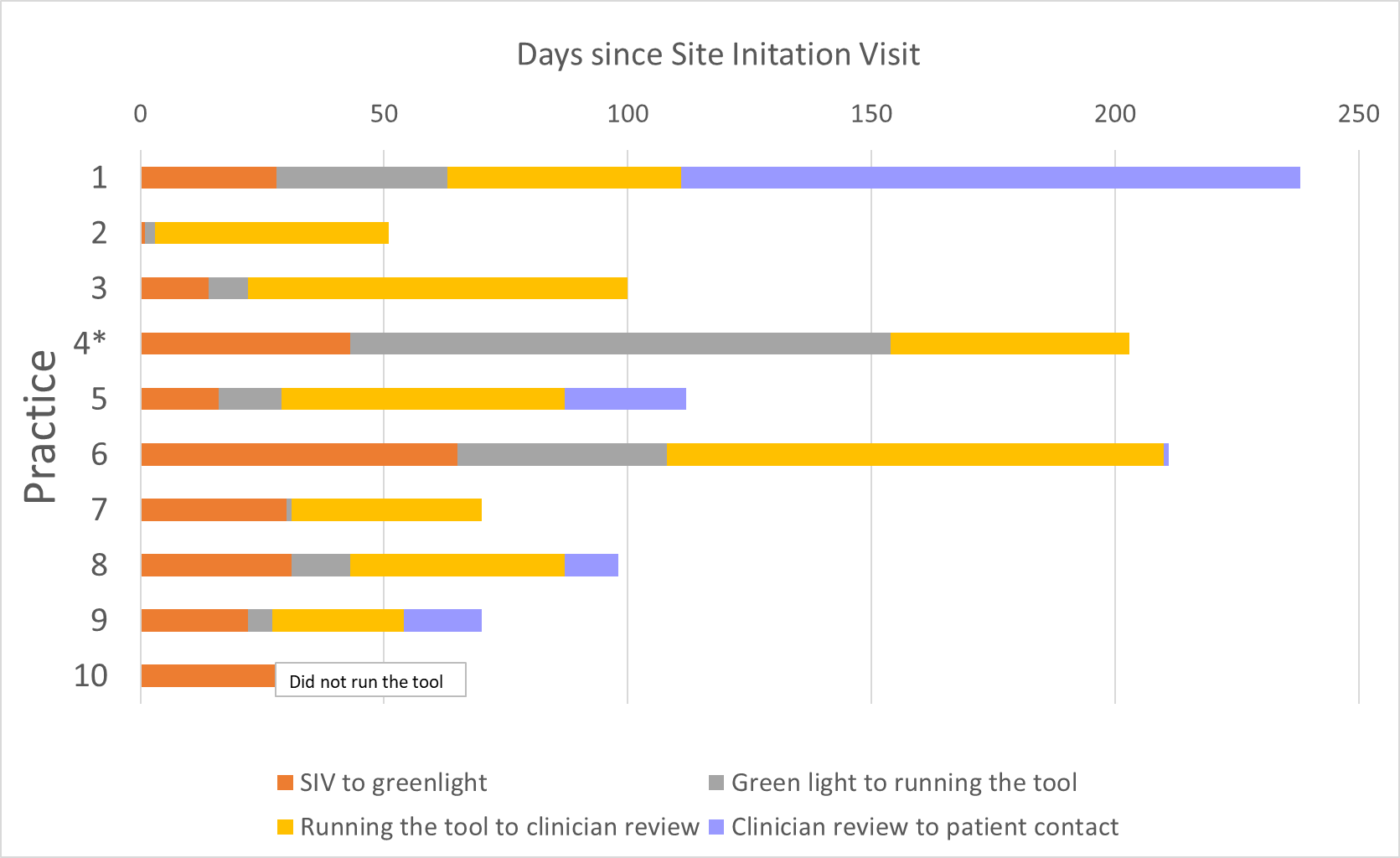

2. National Health Service. National Genomic Test Directory: Testing Criteria for Rare and Inherited Disease. London: NHS, 2026.

3. National Institute for Health and Care Excellence. Nice Guideline NG17: Type 1 diabetes in adults: diagnosis and management. London: NICE, 2015.

4. Jones AG, Hattersley AT. The clinical utility of C-peptide measurement in the care of patients with diabetes. *Diabet Med* 2013;30(7):803-17. doi: 10.1111/dme.12159 [published Online First: 2013/02/19]

5. Bingley PJ. Clinical applications of diabetes antibody testing. *J Clin Endocrinol Metab* 2010;95(1):25-33. doi: 10.1210/jc.2009-1365 [published Online First: 2009/10/31]
